# The burden of dengue and risk factors of transmission in nine districts in Sri Lanka

**DOI:** 10.1101/2023.04.23.23288986

**Authors:** Chandima Jeewandara, Maneshka Vindesh Karunananda, Suranga Fernando, Saubhagya Danasekara, Gamini Jayakody, S. Arulkumaran, N.Y. Samaraweera, Sarathchandra Kumarawansha, Subramaniyam Sivaganesh, P. Geethika Amarasinghe, Chintha Jayasinghe, Dilini Wijesekara, Manonath Bandara Marasinghe, Udari Mambulage, Helanka Wijayatilake, Kasun Senevirathne, A.D.P Bandara, C.P. Gallage, N.R. Colambage, A.A. Thilak Udayasiri, Tharaka Lokumarambage, Y. Upasena, W.P.K.P. Weerasooriya, seroprevalence study group, Graham S. Ogg, Gathsaurie Neelika Malavige

## Abstract

**Background:** It is crucial to understand the differences in dengue seroprevalence rates in different regions in Sri Lanka to understand the burden of infection to plan dengue vaccination programmes.

**Methods:** age stratified seroprevalence rates were assessed in 5208 children, aged 10 to 19 years, in nine districts representing the nine provinces in Sri Lanka. A stratified multi-stage cluster was used to select 146 schools representing each district. Probability proportionate to the size (PPS) sampling technique based on the age distribution of general population and the urbanicity in each district was used to select the number of clusters to be enrolled for the study from each district.

**Findings:** The overall dengue seroprevalence rates in children was 24.8%, with the highest rates reported from Trincomalee (54.3%) and the lowest rates from Badulla (14.2%), which is a high altitude estate area. There was a weak but positive correlation between the dengue antibody positivity rates and age in districts which had seroprevalence rates of >25%, while there was no increase in antibody titres with age in the other districts. While the seroprevalence rates was significantly higher in urban areas (35.8%) compared to rural (23.2%) and estate areas (9.4%), there was no association with seropositivity rates with population density (Spearmans r=-0.01, p=0.98), in each district.

**Interpretation:** The seroprevalence rates in many districts were <25% and the rates were very different to those reported from Colombo. Therefore, it would be important to take into account these differences when rolling out dengue vaccines in Sri Lanka.

**Funding:** We are grateful to the World Health Organization and the UK Medical Research Council for support.

## Introduction

Almost half of the global population is at risk of being infected with the dengue virus (DENV), which is the most rapidly spreading mosquito borne viral infection ^1^. The incidence of symptomatic dengue has markedly in increased in recent years from 505,430 cases in 2000 to 5.2 million in 2019, which is a 10-fold increase ^1^. It is predicted that the burden of dengue will increase in the future, as a result of climate change, rapid urbanization, and population growth ^2,3^. Due to the effect on the health care systems in dengue endemic countries, the WHO named it as one of the top ten threats to global health in 2019 ^4^. However, the incidence of dengue continues to rise causing significant impact on the limited health care resources and economies in dengue endemic countries.

So far, control of dengue outbreaks have solely relied on vector control measures by use of insecticides and eliminating breeding places ^1^. Recently, novel technologies for vector control such as using Wolbachia infected mosquitoes have shown promising results ^5^. Apart from vector control, development of a safe and effective vaccine to prevent dengue or reduce hospitalizations has been a key focus and the first dengue vaccine was approved by the FDA in 2019 ^6^. This first dengue vaccine (CYD-TDV) received approval by several countries and was used for mass vaccination of children in the Philippines ^7^. This vaccine was shown to have lower efficacy rates for DENV-2, despite having induced high neutralizing antibody levels in both baseline seronegative and seropositive individuals for all four serotypes. ^8^. Subsequently it was shown that the vaccine-induced immunity waned with time, and that hospitalizations were higher in seronegative vaccine recipients when they were infected with the DENV, possibly due to antibody dependent enhancement (ADE)^9^. As a result these findings, the WHO recommended that the vaccine only to be used in populations with DENV seroprevalence rates >70% or in those who were known to be seropositive for dengue ^10^.

Currently there is another dengue vaccine, TAK-003 which has received approval from the European Medical Agency and has been registered in several countries for prevention of dengue. A further dengue vaccine (TV003) is currently undergoing phase 3 trials. TAK-003, is a live attenuated vaccine based on the DENV-2 backbone ^11^ and was shown to have an overall efficacy of 62.0% for virologically confirmed dengue and an efficacy rate of 83.6% against hospitalizations at the end of a three year follow up period ^11^. The efficacy rate was highest against DENV2 (91.9%) in baseline seronegative individuals, while there was a reduced efficacy against DENV1 (43.5%) and very limited efficacy against DENV3 (-23.4%) ^11^. The TAK-003 was shown to be 100% effective in prevention hospitalization due to DENV2 and DENV1 (86.6%) in DENV seronegative vaccinees, with no efficacy against DENV3. The efficacy rates were lowest in the seronegative children aged 6 to 11 years ^11^. Unpublished data on the manufacturer’s website states that the vaccine efficacy at 4.5 years was 84% against hospitalization without any safety signals ^12^.

While the TAK-003 appears to be very promising without any safety signals, in order to roll out the vaccine most effectively, it is important to understand the burden of disease in each population, transmission dynamics and age stratified seroprevalence rates to understand the true burden of infection. As dengue is very much under-reported in many countries, the case numbers are unlikely to reflect the true infection rates in a particular population. Furthermore, age stratified seroprevalence rates in certain geographical areas in a particular country, are unlikely to reflect the overall seroprevalence rates, as urbanicity, population density, mobility, temperature, and rainfall affect vector densities and outbreaks ^3,13^. Although many age stratified seroprevalence studies of dengue have been done in the Colombo district in years 2003, 2010, 2013 and 2017, there are no data on dengue seroprevalence rates in any other region of the country ^14-17^. Such data are crucial to understand the differences in dengue transmission rates in different regions in Sri Lanka to understand the risk factors associated with dengue transmission and the burden of infection to plan dengue vaccination campaigns.

In this study, we determined the age stratified dengue seroprevalence in nine districts in Sri Lanka, representing the nine provinces, which include urban, rural and estate areas, with some at sea level and other areas which were at a high altitude. These data are important in order to plan dengue vaccination programs in Sri Lanka.

## Methods

### Study participants

Sri Lanka has nice provinces, with each province having 2 to 4 districts (total of 26 districts). To find the age stratified seroprevalence rates in Sri Lanka, we randomly selected one district from each province. All school children between the age of 10 to 19 years, who were attending public or private schools in Sri Lanka, were included following informed written consent from the parents/guardians and after obtaining assent from the children, during September 2022 to 31^st^ March 2023. Children who study at special needs classes, those who moved into the district from another district within 12 months, children who have contraindications for venepuncture or those who have a diagnosed immune deficiency disorder, a cancer or chronic kidney disease, were excluded from the study.

Blood samples (5ml) were collected by venepuncture by a trained nurse or a medical officer under aseptic conditions according to the universal precautions recommended by the WHO, at the school where the child attends. The samples were centrifuged, serum separated and stored in -80 C freezers, until further use for the assays.

### Ethics statement

Ethics approval was obtained by the Ethics review Committee, University of Sri Jayewardenepura and administrative clearance was obtained by the Ministry of Health, Sri Lanka.

### Sampling technique

A stratified multi-stage cluster sampling method was used to select the schools in each district. A cluster size of 40 students was included from a school to get maximum representation from each class. Accordingly, 146 schools were included in the study throughout the country and a multiple-stage cluster sampling method was used to select eligible students from a selected school. As the population density was different in each district, a probability proportionate to the size (PPS) sampling technique based on the age specific (10 to 19 age group) general population in each district was used to select the number of clusters to be enrolled for the study from each district. As different districts have different urbanicity sectors (urban, rural and estate), the PPS sampling technique was also applied to represent specific resident population of each sector in the district (additional details of sampling techniques, data collection, sample size calculation and case definitions are included in supplementary methods).

### Classification of different areas as urban, rural and estate areas

In order to find out the age stratified infection rates based on urbanicity, we had recruited the children from schools of each district which had representation from urban, rural and estate areas. Derived from the latest census for Sri Lanka ^18^, schools in urban areas were defined as those situated in areas administered by municipal and urban councils. Schools in estate areas were defined as those situated in plantations with an extent of 20 acres or more and with 10 or more resident laborers. Schools situated in areas that are neither urban nor estate were defined as rural. Although we recruited children from both urban and rural areas from all districts, children from estate areas were only recruited in districts where estates were present.

### Assessment of dengue IgG antibodies

The presence of DENV-specific IgG antibodies was assessed by using a commercial assay (PanBio Indirect IgG ELISA). We have used this assay for seroprevalence studies in Sri Lanka previously ^14^. This assay is a semi quantitative assay, and the antibody levels are expressed as PanBio units. PanBio units were calculated according to the manufacturer’s instructions and accordingly, PanBio units of > 11 were considered positive, 9-11 was considered equivocal and

< 9 was considered negative.

### Statistical analysis

GraphPad Prism version 8.3 was used for statistical analysis. As the data were not normally distributed, differences in means were compared using the Mann-Whitney U test (two tailed), and the Kruskal-Wallis test was used to compare the differences of the antibody levels in the different districts, and in urban, rural and estate sectors. Spearman rank order correlation coefficient was used to evaluate the correlation between the age and DENV-specific antibody levels (Panbio Units).

## Results

A total of 5208 children were included in the study from the nine districts in Sri Lanka. The overall seroprevalence rates for each district in shown in Table 1. The highest seroprevalence rates were reported in the Trincomalee district followed by Jaffna, Gampaha and Ratnapura. The lowest seroprevalence rates were seen in Badulla, which is in higher altitudes.

**Table 1:** Overall dengue IgG seropositivity rates in each of the nine districts in Sri Lanka.

| **District** | **Age group**<br>**10 to 12**<br>**N (%)** | **Age group**<br>**13 to 15**<br>**N (%)** | **Age group**<br>**16 to 20**<br>**N (%)** | **All age groups**<br>**N (%)** |
| --- | --- | --- | --- | --- |
| Trincomalee | 40(55.6) | 48(48.5) | 56(66.7) | 144 (54.3) |
| Jaffna | 32(34.0) | 35(36.1) | 49(37.7) | 116 (31.3) |
| Kurunegala | 38(14.5) | 45(16.0) | 45(15.8) | 128 (15.5) |
| Matara | 18(11.5) | 30(17.9) | 34(18.8) | 82 (16.2) |
| Ratnapura | 26(16.7) | 49(29.5) | 74(42.8) | 149 (30.0) |
| Polonnaruwa | 21(33.9) | 24(27.6) | 27(25) | 72 (28.02) |
| Gampaha | 117(27.5) | 142(30.7) | 156(33.4) | 415 (30.6) |
| Kandy | 26(14.1) | 31(13.2) | 59(22.5) | 116 (17.03) |
| Badulla | 18(10.3) | 31(18.0) | 22(14.2) | 71 (14.2) |
| Total | 336(21.0) | 435(24.6) | 522(28.3) | 1293 (24.8) |

### Age stratified seroprevalence rates and FOI in different districts in Sri Lanka

As there is a significant variation in reporting of dengue cases in the country and as we observed marked differences in the overall seroprevalence rates in each district, we proceeded to evaluate the age stratified seroprevalence rates in each district. The age stratified seroprevalence rates of each of the nine districts are shown in supplementary data (Supplementary Tables 1-9). We found that while the seroprevalence rates increased with age in some districts such as Trincomalee, Jaffna, Gampaha and Ratnapura, which reported the highest seropositivity rates, there was no such increase with age in the other districts (Figure 1A).

**Figure 1:**
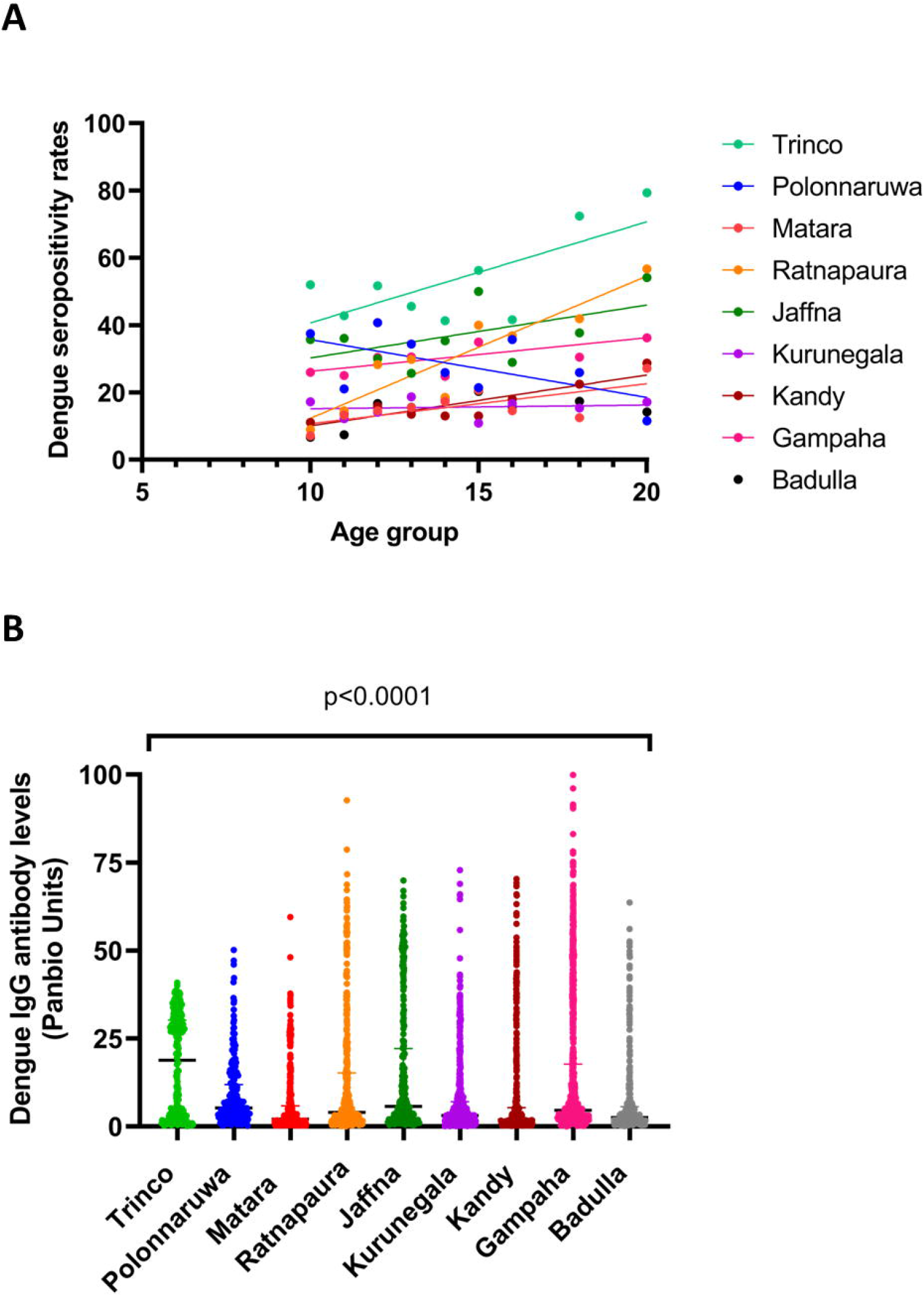
Age stratified seropositivity rates of dengue and DENV-specific IgG levels in nine districts in Sri Lanka. The presence of DENV specific IgG antibodies were measured in 5208, from nine districts in Sri Lanka. The seropositivity rate was calculated for each age group in each district and correlated with age (A). Spearman rank order correlation coefficient was used to evaluate the correlation between the age and seropositivity rates. The presence of DENV specific IgG antibodies were measured in 5208, from nine districts in Sri Lanka using a commercial ELISA and the DENV-specific IgG antibodies semi-quantitively expressed as Panbio units (B). The bars indicate the median and the interquartile range. The Kruskal-Wallis test was used to compare the differences of the antibody levels in the different districts, p<0.0001 (two-tailed).

Trincomalee had the highest dengue seroprevalence rates among all districts and the highest seropositivity rates at 10 years of age (Supplementary Tables 1-9). At 10 years 52% of children were seropositive for dengue, which increased to 72.4% to 79% in 17- to 20-year-olds (Supplementary Tables 1-9). Badulla, which is mainly a rural and estate area with high altitude had the lowest seropositivity rates for dengue. At 10 years of age, only 6.7% of children were seropositive for dengue, which did increase with age, although this was not significant (Spearman’s r=0.43, p=0.25) (Supplementary Tables 1-9).

### Dengue antibody levels of different districts

As repeated infection with the DENVs could lead to higher DENV-specific IgG antibody levels, we compared the DENV antibody levels (Panbio units, which is an indirect measure of envelope protein specific IgG antibodies), in different districts. The median antibody levels were highest in the Trincomalee district (median 18.8, IQR 2.7 to 30.3 Panbio units), which also had the highest seroprevalence rates (Figure 1B). The median DENV-specific IgG levels in Polonnaruwa, Jaffna, Gampaha and Ratnapura districts were similar.

If the transmission intensity is assumed to be constant over time, the rate of infection remains constant and the rate of susceptible individuals acquiring infection and the rate of seropositive individuals possibly acquiring another infection would increase with age. This would give rise to an increase in DENV-specific IgG titres with age ^19^. Therefore, we also correlated the DENV antibody levels (Panbio units) in different age groups in different districts (Supplementary Figures 1-9). Although the DENV-specific IgG antibody titres significantly correlated with age in many districts, this was a very weak correlation (Spearman’s R between 0.06 to 0.29).

### Risk of dengue transmission in relation to urbanicity and population density

Urbanization is associated with a higher density of Aedes mosquitoes and disease transmission. In addition, higher population densities of over 1000 individuals/km^2^ have also shown to associate with increase in transmission of arbovirus infections ^13^. Therefore, we determined the age stratified seroprevalence rates of dengue in children from urban, rural and estate populations. The overall seroprevalence rates were 35.8% in urban areas, 23.2% in rural areas and 9.4% in estate areas (Table 2). Except for a significant positive correlation with age and seropositivity rates in rural areas (Spearman’s r=0.93, p=0.0007), there was no association with age and seropositivity rates in urban or rural areas (Figure 2A).

**Table 2:** Age stratified seroprevalence of dengue in children from urban, rural and estate areas in Sri Lanka.

| **Age**<br>**group** | **Urban** | **Rural** | **Estate** |
| --- | --- | --- | --- |

|  | Total<br>number | Seropositivity<br>rates<br>N (%) | Total<br>number | Seropositivity<br>rates<br>N (%) | Total<br>number | Seropositivity<br>rates<br>N (%) |
| --- | --- | --- | --- | --- | --- | --- |
| 10 | 68 | 24 (35.3) | 419 | 71 (16.9) | 7 | 1 (14.3) |
| 11 | 89 | 27 (30.3) | 446 | 79 (17.7) | 23 | 0 (0.0) |
| 12 | 102 | 41 (40.2) | 422 | 89 (21.1) | 21 | 4 (19.1) |
| 13 | 113 | 34 (30.1) | 454 | 110 (24.2) | 17 | 1 (5.9) |
| 14 | 104 | 31 (29.8) | 449 | 93 (20.7) | 24 | 1 (4.2) |
| 15 | 122 | 47 (38.5) | 460 | 116 (25.2) | 23 | 2 (8.7) |
| 16 | 94 | 33 (35.1) | 463 | 115 (24.8) | 31 | 5 (16.1) |
| 17-18 | 146 | 56 (38.4) | 661 | 168 (25.4) | 24 | 4 (16.7) |
| 19-20 | 41 | 22 (53.7) | 362 | 119 (32.9) | 22 | 0 (0.0) |
| **Total** | 879 | 315 (35.8) | 4136 | 960 (23.2) | 192 | 18 (9.4) |

**Figure 2:**
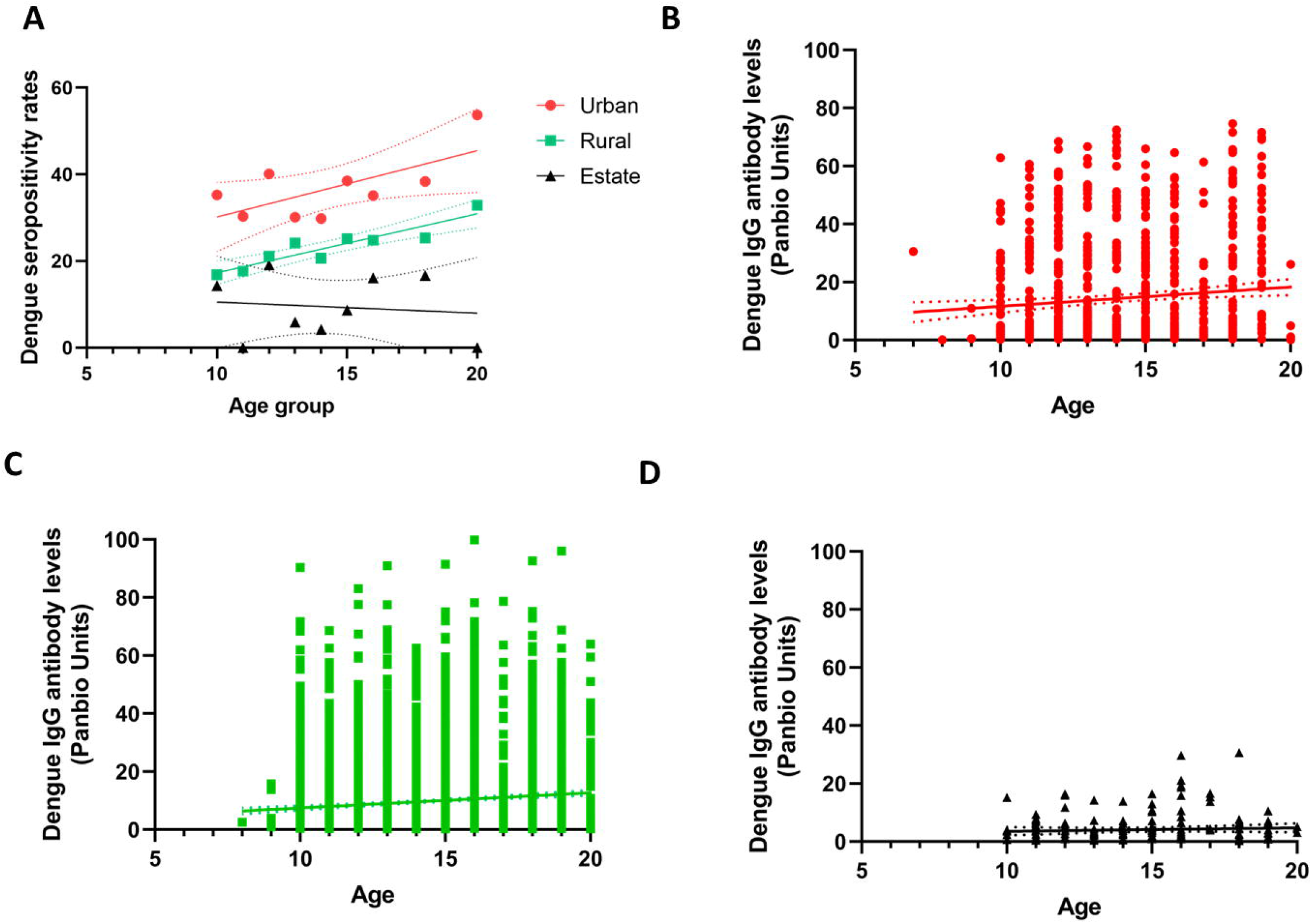
Age stratified seropositivity rates of dengue and DENV-specific IgG levels in urban, rural and estate areas in Sri Lanka. The presence of DENV specific IgG antibodies were measured in urban (n= 888), rural (n=4148) and estate (n=193) areas in Sri Lanka. The seropositivity rate was calculated for each age group in each area and correlated with age (A). Spearman rank order correlation coefficient was used to evaluate the correlation between the age and seropositivity rates. The presence of DENV specific IgG antibodies were measured in urban (B), rural (C) and estate (D) areas using a commercial ELISA and the DENV-specific IgG antibodies semi-quantitively expressed as Panbio units. The DENV-antibody levels were correlated with age in each area.

We investigated the association between DENV-specific IgG levels and age, and although there was a weak but significant correlation between antibody levels (Panbio units) and age in urban (Spearman’s R = 0.11, p=0.0009) and in rural areas (Spearman’s R = 0.11, p<0.0001), there was no correlation in the estate areas (Spearman’s R = 0.06, p=0.38) (Figure 2B, C and D).

As the transmission rates were shown to be higher at higher population densities ^13^, we also sought to determine any association with population density of each district and overall seropositivity rates. We analyzed the association between seropositivity and population density of each district based on population density rates in each district in 2019 ^20^. We did not find any correlation between population density in each district (Spearman’s r=-0.01), p=0.98).

## Discussion

In this study we have assessed the age stratified seroprevalence rates in nine districts in Sri Lanka and investigated the association of seroprevalence rates and antibody titres with urbanicity and population density. We found that the seroprevalence rates were very different in the nine districts in Sri Lanka and markedly different to the seroprevalence rates reported from the Colombo district ^14,15,17^.

Regular outbreaks of dengue occurred in Sri Lanka since 1989, and since then the incidence of dengue has steadily increased ^15^. Although dengue cases are being reported from all over the island, over 50% of cases are being reported in the Western province, with the majority of cases from Colombo. In 2003, Sri Lanka reported a total of 2266 cases and the overall seropositivity rate among children in Colombo was 34% ^16^. In year 2013 when the study was repeated, Sri Lanka reported a total of 41,499 and the seropositivity rate among children was 50.7% ^14^. Although Trincomalee reported dengue outbreaks in recent years, the number of cases are several fold lower than reported in Colombo district and lower than the case number reported from Gampaha, Kandy and many other districts ^21^. Therefore, it is surprising that Trincomalee had the highest seropositivity rates (54.3%), despite lower number of reported cases. This shows that many cases are in fact under reported in many areas. For instance, in year 2022, India reported 110,473 cases for a population of 1.4 billion ^22^, whereas Sri Lanka reported 76,000 cases for a population of 21 million ^21^. However, the age stratified seroprevalence rates and overall seropositivity rates for dengue are higher in many regions in India compared to Sri Lanka^23^.

From 2016 to 2022 (excluding years 2020 and 2021) Sri Lanka has been reporting on an average of 60,000 to 100,000 cases each year (186,101 cases in 2017) ^24^, with an approximate incidence rate of 285.7/100,000 population per year to 428.6/100,000 population per year. The incidence of dengue was several folds lower than reported in many Latin American countries such as Brazil, Honduras, French Guiana, Nicaragua ^25^, India and many Southeast Asian countries such as Thailand, Vietnam and Indonesia ^26^. This is reflected in the lower overall seroprevalence rates of dengue in Sri Lanka and the age stratified seroprevalence rates in many districts compared to many other countries and regions. For instance, 5/9 districts showed seroprevalence rates of <25%, indicating lower transmission rates in these districts. In addition, in these 5 districts with lower seroprevalence rates, the DENV-specific IgG titres did not increase with age, which suggested that they are unlikely to be repeatedly infected with time. Due to the lower seroprevalence rates in many areas in Sri Lanka, the benefits and costs of using a dengue vaccine for prevention of infection dengue in these areas should be carefully assessed. Furthermore, although TAK-003 was shown to have an overall efficacy rate of 83.6% against hospitalizations at the end 3 years, the efficacy rates in seronegative children were negligible for DENV3 ^11^. The vaccine had an overall efficacy of 77.1% against hospitalization in seronegative children and 86% in seropositive children, with an overall efficacy of 65.4% of preventing DHF ^11^. As DENV3 is emerging as the predominant serotype in many countries in Asia^27^ including Sri Lanka (unpublished data from our laboratory), causing massive outbreaks, the cost-benefit and risks of using this vaccine during DENV3 outbreaks should be carefully evaluated. While the vaccine looks quite promising, it would be important to carefully evaluate any long-term safety signals in seronegative children and in populations with low level of transmission.

In this study we also evaluated the factors that could affect seropositivity rates, as the population recruited from each district were from urban, rural and estate areas. We found that the age stratified seroprevalence rates were in fact significantly higher in the urban population, with lower rates in rural areas followed by estate areas, similar to what was observed in India ^23^. Although urbanicity has shown to associate with higher transmission rates, active surveillance for dengue carried out in Cambodia showed a similar dengue incidence in urban and rural areas ^28^. Furthermore, seroprevalence studies in Malaysia have shown that seropositivity rates were similar in urban and rural areas, indicating similar transmission rates ^29^. The reasons for similar infection rates in urban and rural population in Malaysia and Cambodia, and lower infection rates in rural Sri Lanka and India are unknown, but could be due to vector type and abundance, climatic factors and mobility of the population (frequency of visits of rural population to urban areas). However, interestingly we did not find an association with population density of each district with the age stratified seroprevalence rates for dengue.

In summary, we have carried out the first island wide seroprevalence study of dengue in Sri Lanka and show that the age stratified seroprevalence rates vary widely in different areas in Sri Lanka. The seroprevalence rates in many districts were <25% and the rates were very different to those reported from Colombo. Therefore, it would be important to take into account these differences in seroprevalence rates in order to evaluate the roll out of dengue vaccines in Sri Lanka.

## Data Availability

All data produced in the present study are available upon reasonable request to the authors.

