## Supplementary methods for "The burden of dengue and risk factors of transmission in nine districts in Sri Lanka"

**Sampling technique: selection of schools and classes from each school**

To select the clusters to be included from each sector, all type 1 schools (clusters), which were separated as male-only schools, female-only schools and mixed schools, were listed by sector according to alphabetical order. The number of clusters allocated to a sector were then be divided approximately into three and the required number from each school category was selected from the three lists, randomly. This was to ensure that within each cluster a more-or-less equal number of male and female students were included into the sample. Lists of such schools was obtained from the Provincial Ministry of Education in Sri Lanka. Forty eligible students from each school were selected for the final sample.

In the selected schools, Grade 5 to 13 classes were listed, and one class was selected randomly from each school. Once the classes are selected in male-only and female-only schools, the eligible students were listed according to alphabetical order and from this list, four students from each grade were selected randomly using electronic random number selection software. In mixed schools, Grade 5 to 13 classes were listed and one class was selected randomly. Once the classes were selected; the eligible male and female students were listed separately and two students from each class were selected randomly from the male and female lists using electronic random number selection software.

**Sample size calculation:**

The sample size calculation was targeted to determine the proportion of school-going children (among the 10-19 age group) who was previously infected with the dengue virus (DENV). The minimum sample size was calculated considering the expected proportion of already dengue infected school-going children between the age of 10 to 19 years.

The formula for Calculating a Population Proportion with Absolute Precision was used to calculate the sample size with the expected proportion. The standard normal deviate was taken as 1.96 for a confidence level of 95%. The absolute precision required on either side of the prevalence value was taken as 3% as more precise estimates are needed when estimating proportions on highly socially sensitive issues among special population groups like school children. In the absence of pervious literature related to the seroprevalence of dengue among school children in Sri Lanka, in these particular districts, it was decided to consider 50% for calculation of the sample size for this study^1^.

Sample size was calculated using the following formula:

**N	=	Z_α_^2^	x P(1-P)**

**d^2^**

**N** – Minimum sample size required

**Z_(1-α)_ - 1.96** (Z_α_ value corresponding to a confidence interval of 95% and an α error of 0.05)

**P** – Expected seroprevalence among school children between the age of 10 – 19 years

**d -** Absolute precision required on either side of the proportion (Considering the feasibility and accuracy of the estimation, it was decided to have 0.03 as the desired precision level to obtain precise estimates)

Therefore, N =	1.96^2^	x 0.5 x 0.5	= 1067

0.03^2^

N = 1067

A cluster sampling method was used to select the individual sampling units in the social, economic, cultural and feasibility context. In order to minimize the design effect due to “clustering” the following adjustment was made during computation of the final study sample^2^. The required sample size was calculated considering the following formulae:

**Design effect = 1+ (b -1) rho** ^3^

**b** = Average number of responses to the item per cluster or cluster size (40 students)

[A cluster for this national level seroprevalence study was defined as a type 1 public sector school in Sri Lanka. The cluster size (40) was decided on aiming to select four students from each eligible grade]

**roh** = Rate of homogeneity

[A measure of the variability between clusters as compared to the variation within a cluster. This is usually estimated using the results of previous studies of similar design and subject. Usually in community-based studies, the rate of homogeneity ranges from 0.1 to 0.4^3^ . This being a school-based national study using stratified multi-stage cluster sampling, the rate of homogeneity will be composed of the components of variability from all stages of the design. Considering the above facts, for the rate of homogeneity minimum value 0.1 was taken^3^.

Design effect = 1 + (40-1)0.1 = 4.9

Therefore, the design effect value considered for the sample size calculation is 4.9

N = 1067 x 4.9

N = 5,228

Assuming that there could be an expected non-response rate of 10% among the targeted school students, the required sample size was adjusted to compensate for the possible non-response.

Adjusted sample size = 5,228 x 1/(1-0.10)

So, the final sample size will be,

N = 5,809

Accordingly, the number of clusters to be included was calculated to be 146

The cluster size being 40, the study included 5840 study units (146 x 40=5840)
