## Supplementary data for "The burden of dengue and risk factors of transmission in nine districts in Sri Lanka"

**Age stratified dengue seroprevalence in Trincomalee**

| **Age Group** | **Total Number in Each Age Group** | **Seropositivity Rate N (%)** | **Equivocal Rate N (%)** |
| --- | --- | --- | --- |
| 10 | 25 | 13(52%) | 0(0%) |
| 11 | 28 | 12(42.8%) | 1(3.57%) |
| 12 | 29 | 15(51.7%) | 1(3.44%) |
| 13 | 31 | 14(45.61%) | 1(3.22%) |
| 14 | 29 | 12(41.3%) | 0(0%) |
| 15 | 39 | 22(56.4%) | 0(0%) |
| 16 | 26 | 12(46.1%) | 0(0%) |
| 17-18 | 29 | 21(72.4%) | 1(3.44%) |
| 19-20 | 29 | 23(79.3%) | 0(0%) |
| Total | 265 | 144(54.3%) | 4(1.5%) |

**Table 1: Number of children in each age group and dengue IgG seropositivity rates in Trinco**

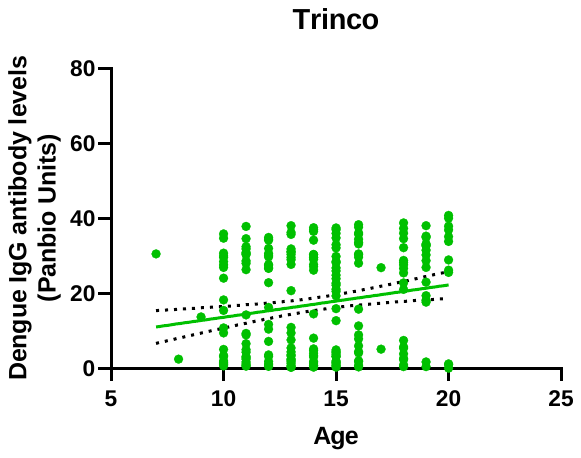

**Figure 1: The association between dengue IgG antibody levels (Panbio titres) and the age of children in Trinco district**

**Age stratified dengue seroprevalence in Polonnaruwa**

| **Age Group** | **Total Number in Each Age Group** | **Seropositivity Rate N (%)** | **Equivocal Rate N (%)** |
| --- | --- | --- | --- |
| 10 | 16 | 6(37.5%) | 1(6.25%) |
| 11 | 19 | 4(21.05%) | 1(5.26%) |
| 12 | 27 | 11(40.74%) | 2(7.41%) |
| 13 | 32 | 11(34.38%) | 1(3.13%) |
| 14 | 27 | 7(25.93%) | 0(0%) |
| 15 | 28 | 6(21.43%) | 3(10.71%) |
| 16 | 28 | 10(35.71%) | 3(10.71%) |
| 17-18 | 54 | 14(25.93%) | 0(0%) |
| 19-20 | 26 | 3(11.54%) | 1(3.85%) |
| **Total** | **257** | **72(28.02%)** | **12(4.67%)** |

**Table 2: Number of children in each age group and dengue IgG seropositivity rates in Polonnaruwa**

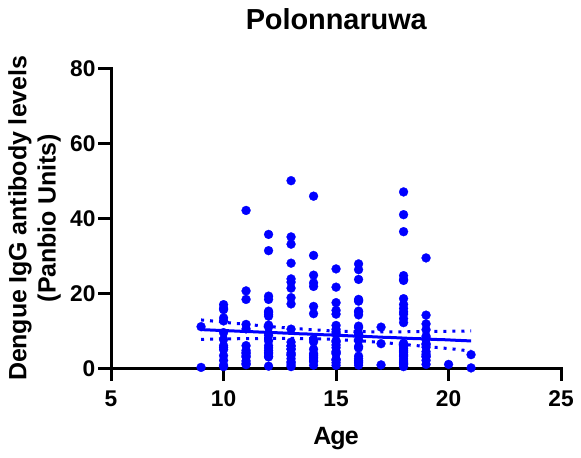

**Figure 2: The association between dengue IgG antibody levels (Panbio titres) and the age of children in Polonnaruwa district**

**Age stratified dengue seroprevalence in Matara**

| **Age Group** |  | **Total Number in Each Age Group** | **Seropositivity Rate N (%)** | **Equivocal Rate N (%)** |
| --- | --- | --- | --- | --- |
| 10 |  | 56 | 4 (7.14%) | 0 (0%) |
| 11 |  | 53 | 7 (13.21%) | 0 (0%) |
| 12 |  | 48 | 7 (14.58%) | 2 (4.17%) |
| 13 |  | 58 | 9 (15.52%) | 1 (1.72%) |
| 14 |  | 52 | 9 (17.31%) | 4 (7.69%) |
| 15 |  | 58 | 12 (20.69%) | 2 (3.45%) |
| 16 |  | 55 | 8 (14.55%) | 2 (3.64%) |
| 17-18 |  | 56 | 7(12.5%) | 4 (7.14%) |
| 19-20 |  | 70 | 19 (27.14%) | 2 (2.86%) |
| Total |  | 506 | 82 (16.21%) | 17 (3.36%) |

**Table 3: Number of children in each age group and dengue IgG seropositivity rates in Matara**

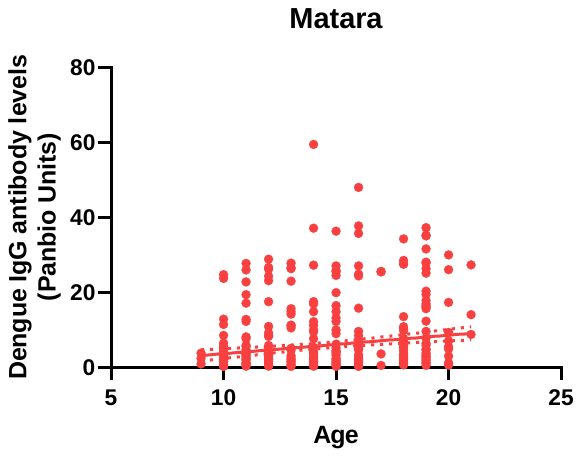

**Figure 3: The association between dengue IgG antibody levels (Panbio titres) and the age of children in Matara district**

**Age stratified dengue seroprevalence in Ratnapura**

| **Age Group** | **Total Number in Each Age Group** | **Seropositivity Rate N (%)** | **Equivocal Rate N (%)** |
| --- | --- | --- | --- |
| 10 | 56 | 5(8.93%) | 1(0.02%) |
| 11 | 54 | 8(14.81%) | 1(1.85%) |
| 12 | 46 | 13(28.26%) | 1(2.17%) |
| 13 | 57 | 17(29.82%) | 1(1.75%) |
| 14 | 54 | 10(18.52%) | 2(3.7%) |
| 15 | 55 | 22(40%) | 2(3.64%) |
| 16 | 57 | 21(36.84%) | 1(1.75%) |
| 17-18 | 86 | 36(41.86%) | 3(3.49%) |
| 19-20 | 30 | 17(56.67%) | 1(3.33%) |
| Total | **495** | **149(30.1%)** | **13(2.63%)** |

**Table 4: Number of children in each age group and dengue IgG seropositivity rates in Ratnapura**

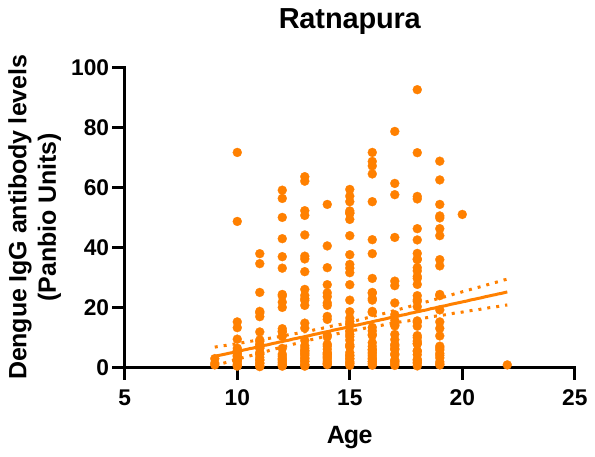

**Figure 4: The association between dengue IgG antibody levels (Panbio titres) and the age of children in Ratnapura district**

**Age stratified dengue seroprevalence in Jaffna**

| **Age Group** | **Total Number in Each Age Group** | **Seropositivity Rate N (%)** | **Equivocal Rate N (%)** |
| --- | --- | --- | --- |
| 10 | 28 | 10 (35.71%) | 2(7.14%) |
| 11 | 36 | 13 (36.11%) | 1(2.78%) |
| 12 | 30 | 9 (30%) | 2(6.67%) |
| 13 | 35 | 9 (25.71%) | 1(2.86%) |
| 14 | 34 | 12 (35.29%) | 1(2.94%) |
| 15 | 28 | 14 (50%) | 0(0%) |
| 16 | 45 | 13 (28.89%) | 1(2.22%) |
| 17-18 | 61 | 23 (37.70%) | 2(3.28%) |
| 19-20 | 24 | 13 (54.17%) | 0(0%) |
| Total | 321 | 116 (36.14%) | 10 (3.12%) |

**Table 5: Number of children in each age group and dengue IgG seropositivity rates in Jaffna**

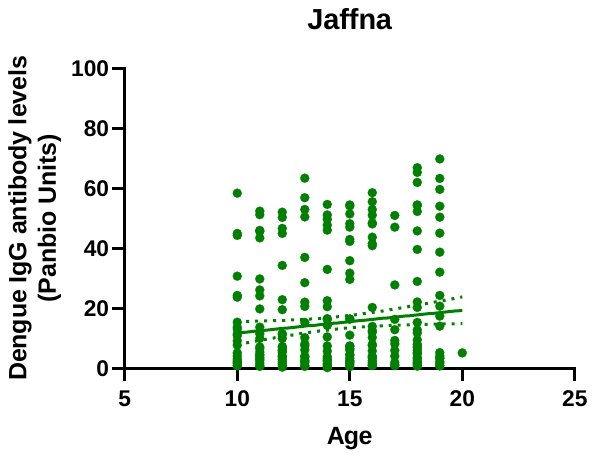

**Figure 5: The association between dengue IgG antibody levels (Panbio titres) and the age of children in Jaffna district**

**Age stratified dengue seroprevalence in Kurunegala**

| **Age Group** | **Total Number in Each Age Group** | **Seropositivity Rate N (%)** | **Equivocal Rate N (%)** |
| --- | --- | --- | --- |
| 10 | 87 | 15 (17.24%) | 1(1.15%) |
| 11 | 90 | 11 (12.22%) | 3(3.33%) |
| 12 | 85 | 12 (14.11%) | 6(7.06%) |
| 13 | 96 | 18 (18.75%) | 3(3.13%) |
| 14 | 93 | 17 (18.27%) | 2(2.15%) |
| 15 | 92 | 10 (10.87%) | 2(2.17%) |
| 16 | 84 | 13 (15.48%) | 1(1.19%) |
| 17-18 | 130 | 20 (15.38%) | 3(2.31%) |
| 19-20 | 70 | 12 (17.14%) | 0(0%) |
| Total | 827 | 128 (15.48%) | 21(2.54%) |

**Table 6: Number of children in each age group and dengue IgG seropositivity rates in Kurunegala**

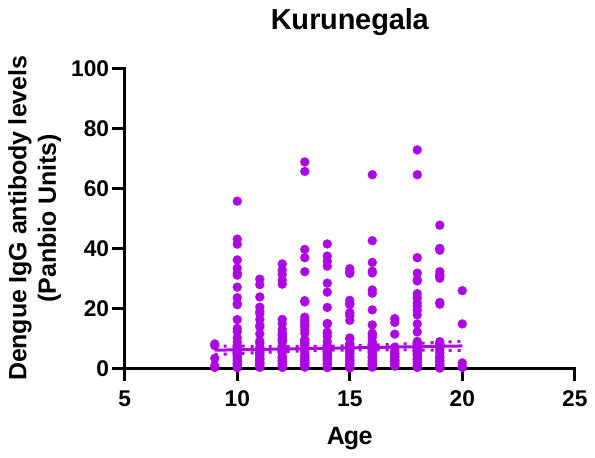

**Figure 6: The association between dengue IgG antibody levels (Panbio titres) and the age of children in Kurunegala district**

**Age stratified dengue seroprevalence in Kandy**

| **Age Group** | **Total Number in Each Age Group** | **Seropositivity Rate N (%)** | **Equivocal Rate N (%)** |
| --- | --- | --- | --- |
| 10 | 36 | 4 (11.11%) | 0 (0%) |
| 11 | 80 | 11 (13.75%) | 1 (1.25%) |
| 12 | 69 | 11 (15.94%) | 2 (2.9%) |
| 13 | 74 | 10 (13.51%) | 1 (1.35%) |
| 14 | 76 | 10 (13.15%) | 1 (1.32%) |
| 15 | 84 | 11 (13.01%) | 0 (0%) |
| 16 | 100 | 18 (18%) | 3 (3%) |
| 17-18 | 89 | 20 (22.47%) | 0 (0%) |
| 19-20 | 73 | 21 (28.77%) | 2 (2.73%) |
| Total | 681 | 116 (17.03%) | 10 (1.47%) |

**Table 7: Number of children in each age group and dengue IgG seropositivity rates in Kandy**

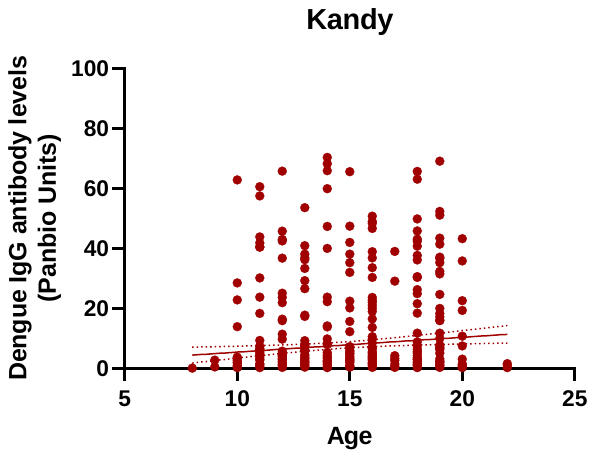

**Figure 7: The association between dengue IgG antibody levels (Panbio titres) and the age of children in Kandy district**

**Age stratified dengue seroprevalence in Gampaha**

| **Age Group** | **Total Number in Each Age Group** | **Seropositivity Rate N (%)** | **Equivocal Rate N (%)** |
| --- | --- | --- | --- |
| 10 | 130 | 35(26%) | 5(3.84%) |
| 11 | 144 | 36(25%) | 5(3.47%) |
| 12 | 151 | 46(30.46%) | 3(1.98%) |
| 13 | 156 | 50(32.05%) | 6(3.84%) |
| 14 | 149 | 37(24.83%) | 5(3.35%) |
| 15 | 157 | 55(35%) | 5(3.18%) |
| 16 | 144 | 53(36.8%) | 2(1.38%) |
| 17-18 | 243 | 74(30.45%) | 5(2.0%) |
| 19-20 | 80 | 29(36.25%) | 6(7.5%) |
| Total | 1354 | 415(30.64%) | 42(3.1%) |

**Table 8: Number of children in each age group and dengue IgG seropositivity rates in Gampaha**

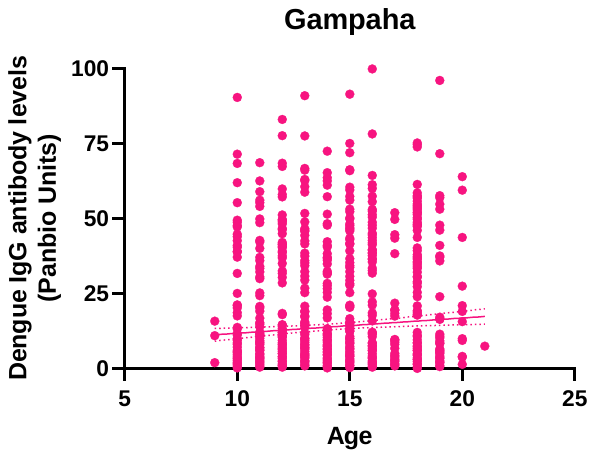

**Figure 8: The association between dengue IgG antibody levels (Panbio titres) and the age of children in Gampaha district**

**Age stratified dengue seroprevalence in Badulla**

| **Age Group** | **Total Number in Each Age Group** | **Seropositivity Rate N (%)** | **Equivocal Rate N (%)** |
| --- | --- | --- | --- |
| 10 | 60 | 4 (6.67%) | 0 (0%) |
| 11 | 54 | 4 (7.41%) | 0 (0%) |
| 12 | 60 | 10 (16.67%) | 2 (3.33%) |
| 13 | 45 | 7 (15.56%) | 1 (2.22%) |
| 14 | 63 | 11 (17.46%) | 1 (1.59%) |
| 15 | 64 | 13 (20.31%) | 3 (4.69%) |
| 16 | 49 | 5 (10.20%) | 1 (2.04%) |
| 17-18 | 83 | 13 (15.66%) | 2 (2.41%) |
| 19-20 | 23 | 4 (17.39%) | 1 (4.35%) |
| Total | 501 | 71 (14.17%) | 11 (2.20%) |

**Table 9: Number of children in each age group and dengue IgG seropositivity rates in Badulla**

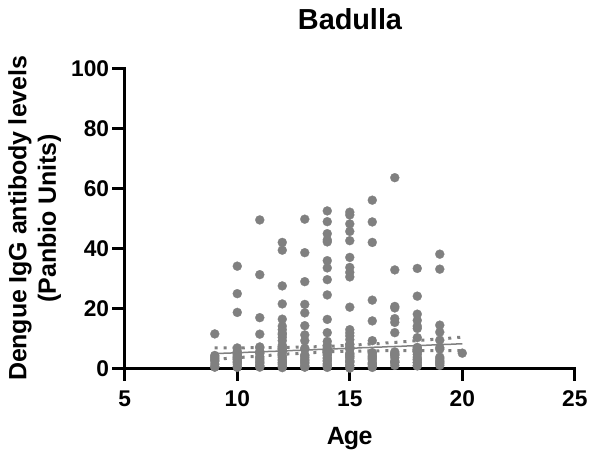

**Figure 9: The association between dengue IgG antibody levels (Panbio titres) and the age of children in Badulla district**
